# Anakinra To Prevent Respiratory Failure In COVID-19

**DOI:** 10.1101/2020.10.28.20217455

**Authors:** Evdoxia Kyriazopoulou, Periklis Panagopoulos, Simeon Metallidis, George N. Dalekos, Garyfallia Poulakou, Nikolaos Gatselis, Eleni Karakike, Maria Saridaki, Georgia Loli, Aggelos Stefos, Danai Prasianaki, Sarah Georgiadou, Olga Tsachouridou, Vasileios Petrakis, Konstantinos Tsiakos, Maria Kosmidou, Vassiliki Lygoura, Maria Dareioti, Haralampos Milionis, Ilias C. Papanikolaou, Karolina Akinosoglou, Dimitra-Melia Myrodia, Areti Gravvani, Aliki Stamou, Theologia Gkavogianni, Konstantina Katrini, Theodoros Marantos, Ioannis P. Trontzas, Konstantinos Syrigos, Loukas Chatzis, Stamatios Chatzis, Nikolaos Vechlidis, Christina Avgoustou, Stamatios Chalvatzis, Miltiades Kyprianou, Jos W. M. van der Meer, Jesper Eugen-Olsen, Mihai G. Netea, Evangelos J. Giamarellos-Bourboulis

## Abstract

**Introduction:** The management of pneumonia caused by SARS-CoV-2 should rely on early recognition of the risk for progression to severe respiratory failure (SRF) and its prevention. We investigated if early suPAR (soluble urokinase plasminogen activator receptor)-guided anakinra treatment could prevent COVID-19-assocated SRF.

**Methods:** In this open-label prospective trial, 130 patients admitted with SARS-CoV-2 pneumonia SARS-CoV-2 and suPAR levels ≥6 μg/l were assigned to subcutaneous anakinra 100mg once daily for 10 days. The primary outcome was the incidence of SRF at day 14. Secondary outcomes were 30-day mortality, changes in sequential organ failure assessment (SOFA) score, of cytokine-stimulation pattern and of circulating inflammatory mediators. Equal number of propensity score-matched comparators for comorbidities, severity on admission and standard-of care (SOC) were studied.

**Results:** The incidence of SRF was 22.3% (95% CI, 16.0-30.2%) among anakinra-treated patients and 59.2% (95% CI, 50.6-67.3%; P: 4.6 x 10^−8^) among SOC comparators (hazard ratio, 0.30; 95%CI, 0.20-0.46). 30-day mortality was 11.5% (95% CI, 7.1-18.2%) and 22.3% (95% CI, 16.0-30.2%) respectively (hazard ratio 0.49; 95% CI 0.25-0.97%; P: 0.041). Anakinra treatment was associated with decrease in SOFA score and in circulating interleukin (IL)-6, sCD163 and sIL2-R; the serum IL-10/IL-6 ratio on day 7 was inversely associated with the change in SOFA score. Duration of stay at the intensive care unit and at hospital was shortened compared to the SOC group; the cost of hospitalization was decreased.

**Conclusions:** Early suPAR-guided anakinra treatment is associated with decrease of the risk for SRF and restoration of the pro- /anti-inflammatory balance.

**Trial Registration:** ClinicalTrials.gov, NCT04357366

## INTRODUCTION

Severe infection by the novel coronavirus SARS-CoV-2 (known as COVID-19) is associated with complex immune dysregulation of the host and it is usually accompanied by unfavourable outcome^1^. When severe respiratory failure (SRF) necessitating mechanical ventilation (MV) emerges, two separate immune phenomena predominate in the infected host; i) macrophage activation syndrome; or ii) complex immune dysregulation with down-regulation of the human leukocyte antigen DR on circulating monocytes, lymphopenia and over-production of interleukin (IL)-6 by monocytes^1^.

It is hypothesized that these immune reactions start early in patients with lower respiratory tract infection (LRTI) by SARS-CoV-2 and are progressively enhanced so as to lead to SRF. This has been suggested to be due to the early release of IL-1 from the lung epithelial cells that are infected by the virus; IL-1 stimulates further cytokine production from alveolar macrophages^2^.

As a consequence, it is assumed that early start of anti-IL-1 anti-inflammatory treatment may prevent SRF^3^. However, most probably not all patients with LRTI by SARS-CoV-2 are in need of early treatment and a screening tool to capture those who are likely to progress to SRF would be an asset. Soluble urokinase plasminogen activator receptor (suPAR) seems to be such a screening tool. We and others, recently demonstrated that suPAR concentrations above 6 μg/l herald worsening to SRF 14 days earlier^4,5^. The positive predictive value for the early prediction of SRF was as high as 85.9%. uPAR is anchored to the cell membranes of the lung endothelial cells. As result of the activation of kallikrein, uPAR is cleaved and enters the systemic circulation as the soluble counterpart suPAR^6^.

We conducted the single-arm open label SAVE trial (<u>s</u>uPAR-guided <u>A</u>nakinra treatment for <u>V</u>alidation of the risk and <u>E</u>arly management of severe respiratory failure by COVID-19) to investigate whether the early administration of anakinra in patients with LRTI due to SARS-CoV-2 and suPAR equal to or greater than 6 μg/l, may prevent the development of SRF. Anakinra is the recombinant soluble IL-1 receptor antagonist that blocks both IL-1α and IL-1β. We report herein the results of the interim analysis in the first 130 enrolled patients and compare the efficacy of anakinra with patients receiving standard-of-care (SOC) treatment.

## METHODS

### Trial oversight

SAVE is an ongoing open-label non-randomized trial conducted in six study sites in Greece (EudraCT number 2020-001466-11; National Ethics Committee approval 38/20; National Organization for Medicines approval ISO 28/20; ClinicalTrials.gov registration NCT04357366). Comparators receiving SOC treatment were hospitalized at the same time period in eight departments of Internal Medicine in tertiary hospitals of Athens who were participating in the registry of the Hellenic Sepsis Study Group without participating in the SAVE trial (www.sepsis.gr). The trial was conducted by the Hellenic Institute for the Study of Sepsis (HISS) and funded in part by HISS, by Technomar Shipping Inc, by Swedish Orphan Biovitrum AB and by the Horizon 2020 RISKinCOVID grant. The funders had no role in the design, conduct, analysis and interpretation of data, and decision to publish. The laboratory of Immunology of Infectious Diseases of the 4^th^ Department of Internal Medicine at ATTIKON University General Hospital served as a central laboratory for the study. The initial draft of the manuscript was written by the first and the last author. All authors vouch for the adherence of the trial to the protocol and first and last author vouch for the accuracy and completeness of the data and analysis.

### Patients

Enrolled patients were adults hospitalized with confirmed infection by SARS-CoV-2 virus by real-time PCR reaction of nasopharyngeal secretions; radiological findings compatible with LRTI; and plasma suPAR level ≥ 6 μg/l using the suPARnostic® Quick Triage kit (Virogates S/A, Blokken 45, 3460 Birkerød, Denmark). Exclusion criteria were: any stage IV malignancy; any do not resuscitate decision; ratio of partial oxygen pressure to the fraction of inspired oxygen (pO_2_/FiO_2_) less than 150; need for MV or non-invasive ventilation under positive pressure (NIV); any primary immunodeficiency; neutropenia (<1,500/mm^3^); any intake of corticosteroids at a daily dose ≥ 0.4mg/kg prednisone or equivalent the last 15 days; any anti-cytokine biological treatment the last month; and pregnancy or lactation. SOC comparators were meeting the same inclusion criteria and did not meet any of the exclusion criteria of the SAVE trial. Written informed consent was provided by the patient or legal representative before screening.

### Trial interventions

Enrolled patients received 100mg anakinra subcutaneously once daily for 10 days. All other drugs were allowed. Fifteen ml of whole blood was collected before start and seven days after start of anakinra and collected into EDTA-coated tubes and sterile and pyrogen-free tubes for the isolation of peripheral blood mononuclear cells (PBMCs), serum and plasma. PBMCs were stimulated for cytokine production. Blood was sampled from SOC comparators on the day of hospital admission and repeated after seven days; plasma and serum were prepared. Biomarkers and cytokines were measured in plasma, serum and supernatants of PBMC cultures. The cost of hospitalization was also provided.

### Outcomes

The incidence of SRF until day 14 was the primary outcome. SRF was defined as any decrease of pO_2_/FiO_2_ below 150 necessitating MV or NIV. Patients dying before day 14 were considered meeting the primary endpoint. Secondary outcomes were 30-day mortality; and the changes of respiratory symptoms score, of sequential organ failure assessment (SOFA) score, of PBMC cytokine stimulation and of circulating plasma inflammatory mediators between days 1 and 7. Stay in the intensive care unit (ICU) for patients who would be in need of MV, total hospital stay and the cost of hospitalization were exploratory endpoints.

Adverse Events (AE) (Common Terminology Criteria for Adverse Events, version 4.03) and Serious Adverse Events (SAE) (see Supplementary Appendix) were captured.

### Statistical analysis

The sample size was calculated assuming the incidence of SRF would decrease from 60% to 45% with anakinra treatment. To achieve so with 90% power at the 5% level of significance, 260 patients were needed. An interim analysis was planned when the first 130 patients would be enrolled.

Qualitative data were presented as percentages with confidence intervals (CI) and quantitative data as median with quartiles. Among all selected comparators receiving SOC treatment, 130 comparators were selected by propensity score 1:1 matching with patients treated with anakinra with SOC. Matching criteria were: age; Charlson’s comorbidity index (CCI); admission severity scores namely pneumonia severity index (PSI), acute physiology and chronic health evaluation (APACHE) II score, SOFA score and WHO severity classification of COVID-19; and SOC treatment with azithromycin, hydroxychloroquine and dexamethasone. Comparison with anakinra-treated patients was performed by the Fisher’s exact test using confirmatory forward stepwise Cox analysis (IBM SPSS Statistics v. 25.0). Comparisons of cytokines and mediators between groups were done by the Student’s “t-test” for parametric variables; and by the Mann-Whitney U test for non-parametric variables. Paired comparisons were done by the Wilcoxon’s rank-signed test. Non-parametric correlations were done according to Spearman. Any two-sided P value <0.05 was statistically significant.

## RESULTS

### Trial conduct

Interim analysis was performed when the 30-day follow-up of the first 130 patients was completed. The first patient was enrolled on April 16^th^ 2020 and the last on September 12^th^ 2020; the inclusion of SOC comparators was done within the same time frame. After propensity-matching, 130 SOC comparators were selected. The study flow chart is shown in Figure S1. 211 patients were excluded because they had suPAR less than 6 μg/l; patients were followed-up and SRF developed in only one (0.47%) patient. Baseline demographics of patients receiving anakinra with SOC treatment and of patients receiving only SOC treatment are shown in Table 1; baseline demographics did not differ.

**Table 1.**
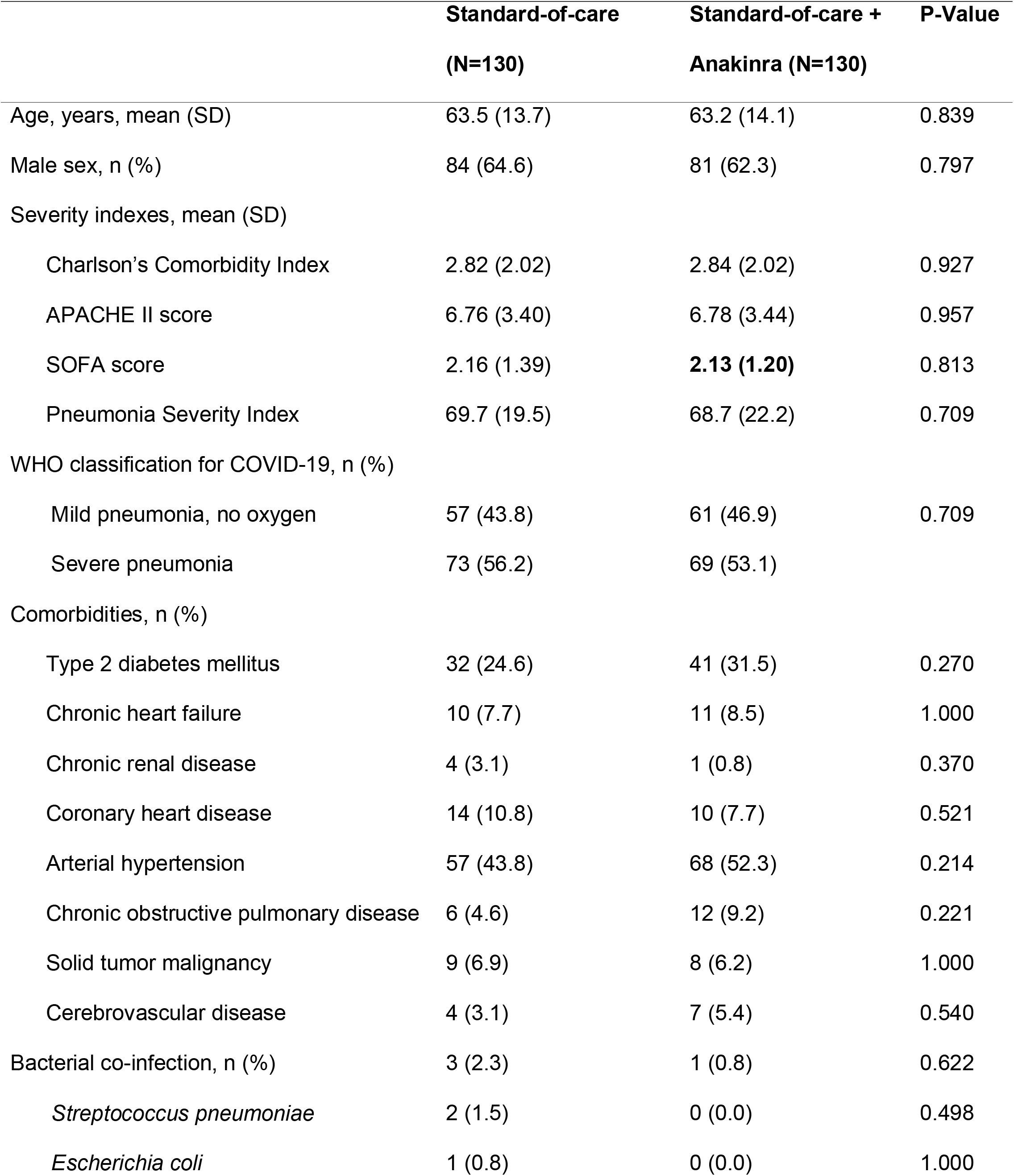
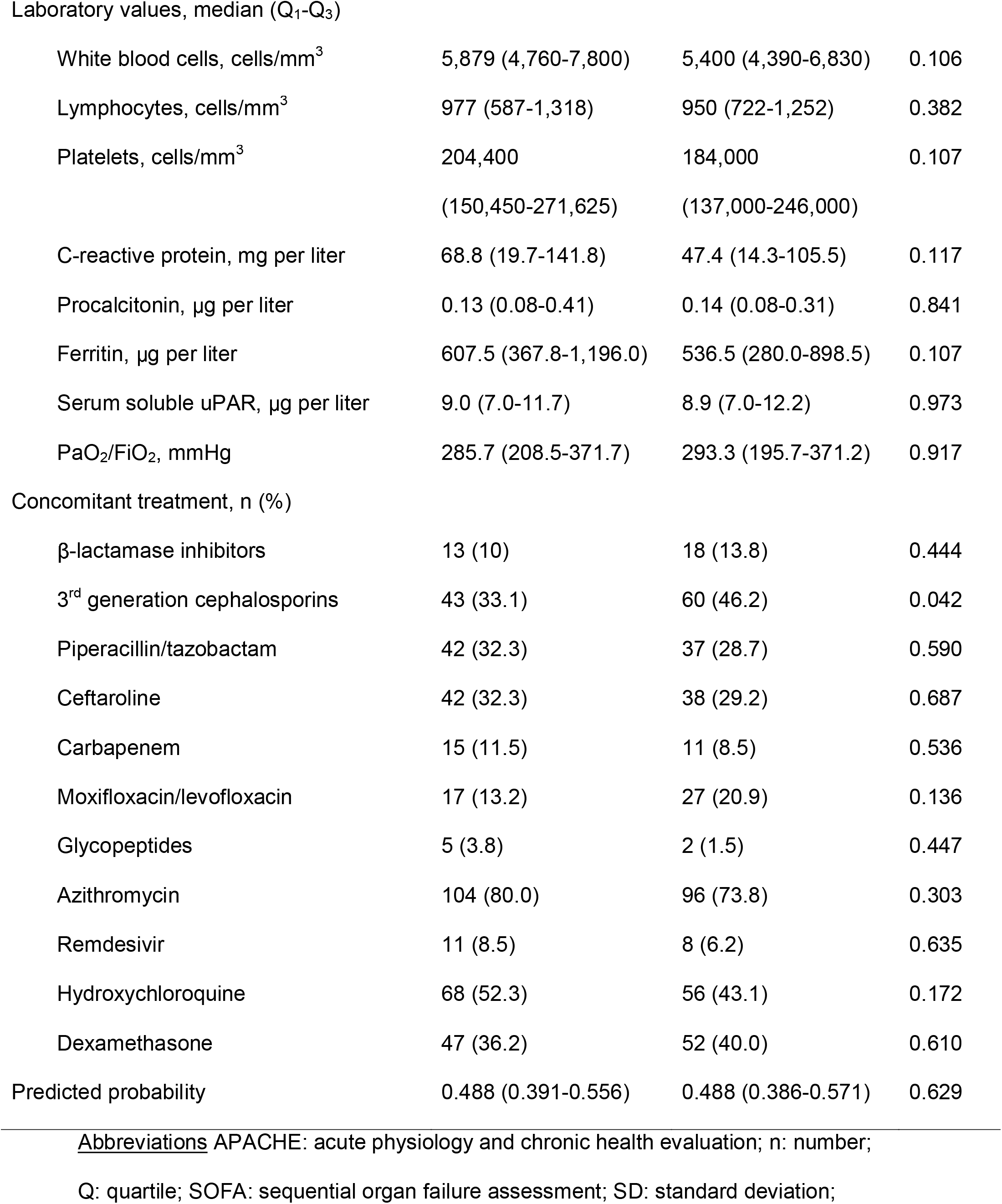
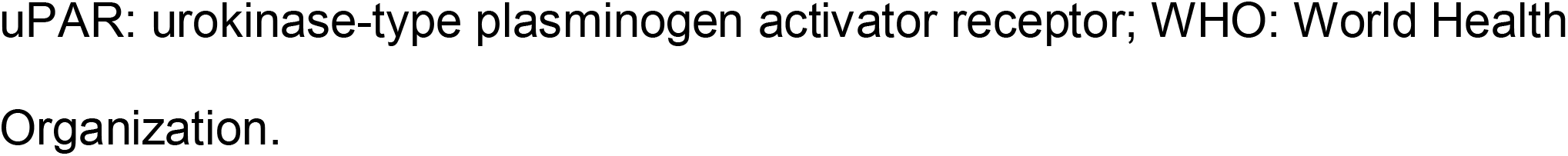
Baseline Characteristics of Patients.

### Study endpoints

Twenty-nine patients (22.3%; 95% CI, 16.0-30.2%) among the intention-to-treat population with anakinra and SOC treatment progressed to SRF until day 14. The incidence of SRF among the 130 SOC-treated comparators was 59.2% (n= 77) (95% CI, 50.6-67.3% P: 4.6 x 10^−8^) (hazard ratio 0.30; 95%CI, 0.20-0.46) (Figure 1A). Multivariate step-wise Cox regression analysis for variables showed that anakinra treatment was the only independent variable protective from SRF (hazard ratio 0.28; 95% CI, 0.18-0.44; P: 2.4 × 10^−8^) (Table S1). One separate multivariate step-wise Cox regression analysis among patients treated with dexamethasone showed anakinra to be the only independent variable protective from SRF (Table S2).

**Figure 1.**
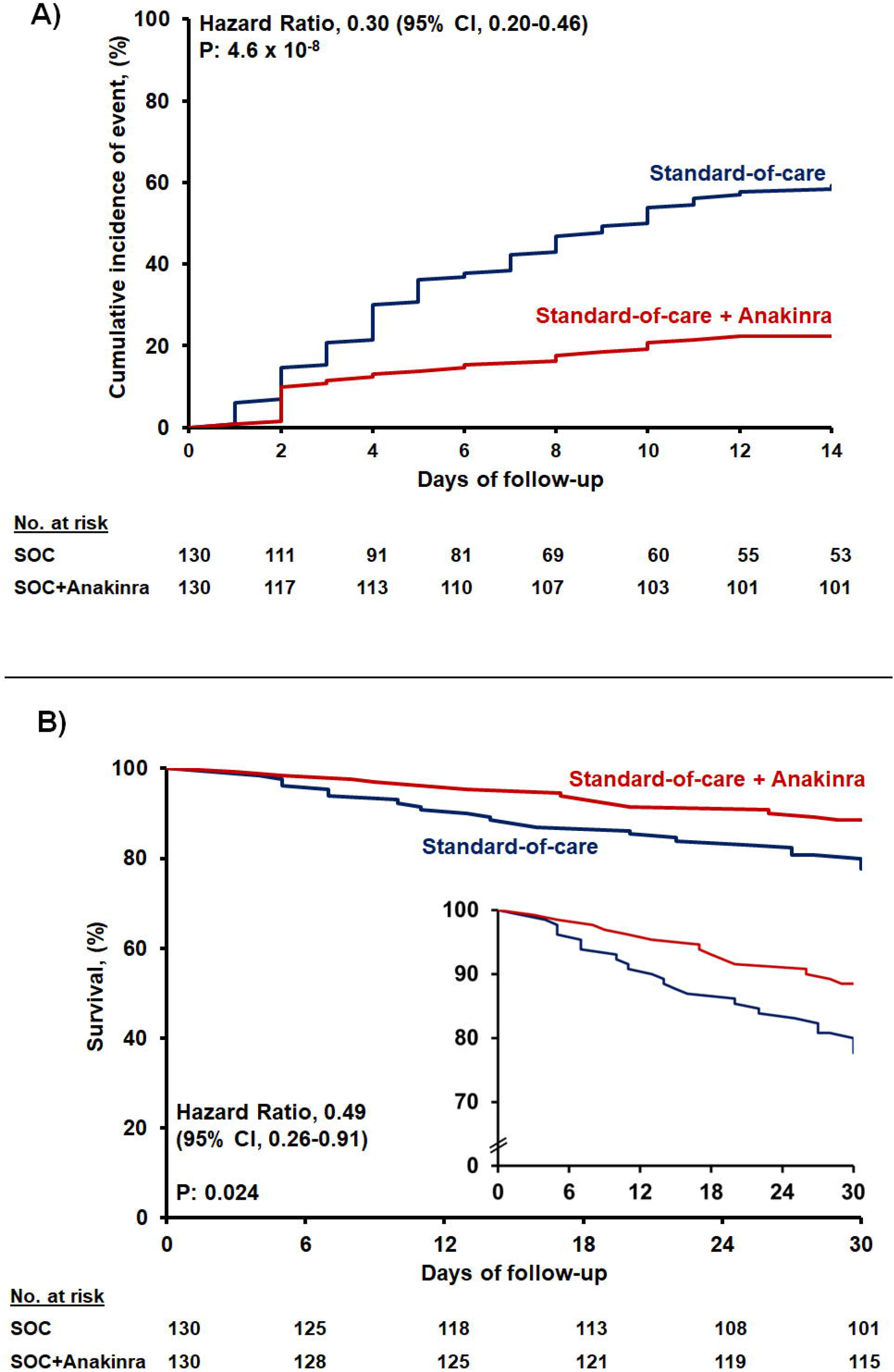
Outcome of patients treated with anakinra. A)Curves of the cumulative incidence of severe respiratory failure (SRF); and B) curves of 30-day mortality. <u>Abbreviations</u> CI: confidence intervals; SOC: standard-of-care

Anakinra treatment was of benefit in most of secondary study endpoints, i.e. 30-day mortality; absolute change of SOFA score by day 14; and absolute change of the respiratory symptoms score by days 7 and 14 (Table 2). Mortality of anakinra treatment after 30 days was 11.5% (95% CI, 7.1-18.2%); this was 22.3% (95% CI, 16.0-30.2%) in patients receiving SOC (Figure 1B). Multivariate step-wise Cox regression analysis for variables showed that anakinra treatment was the only independent variable protective from 30-day mortality (hazard ratio 0.49; 95% CI 0.25-0.97; P: 0.041) (Table S3).

**Table 2.** Primary and Secondary Study Outcomes.

|  | Standard-of-care (N=130) | Standard-of-care +<br>Anakinra (N=130) | OR (95% CI) | P Value |
| --- | --- | --- | --- | --- |
| Severe Respiratory Failure by day 14, n (%) | 77 (59.2) | 29 (22.3) | 0.19 (0.12-0.34) | **1.8 x 10<sup>-9</sup>** |
| Mechanical Ventilation by day 14, n (%) | 65 (50.0) | 25 (19.2) | 0.24 (0.14-0.42) | **2.7 x 10<sup>-7</sup>** |
| Non-invasive Mechanical Ventilation by day 14, n (%) | 9 (6.9) | 6 (4.6) | 0.65 (0.23-1.88) | 0.596 |
| 14-day mortality, n (%) | 16 (12.3) | 6 (4.6) | 0.35 (0.13-0.91) | **0.043** |
| Absolute change of SOFA score by Day 7 compared to<br>baseline, median (Q1 to Q3) | 0 (-1 to 1) | 0 (-1 to 0) | NA | 0.356 |
| Absolute change of SOFA score by Day 14 compared to<br>baseline, median (Q1 to Q3) | 0 (-1 to 6) | -2 (-1 to 0) | NA | **0.004** |
| Absolute change of respiratory symptoms score by Day 7<br>compared to baseline, median (Q1to Q3) | 0 (0 to 0.75) | -1 (-3 to 0) | NA | **0.019** |
| Absolute change of respiratory symptoms score by Day 14<br>compared to baseline, median (Q1 to Q3) | 0 (-0.75 to 0) | -2 (-4 to -1) | NA | **0.016** |
| 30-day mortality, n (%) | 29 (22.3) | 15 (11.5) | 0.45 (0.23-0.90) | **0.031** |

Abbreviations: CI: confidence interval; n: number; NA: non-applicable; OR: odds ratio; Q: quartile; SOFA: sequential organ failure assessment

Two main secondary study outcomes were the effects of anakinra treatment on circulating inflammatory biomarkers and on the function of PBMCs. Compared to SOC comparators, anakinra-treated subjects experienced increase of the absolute lymphocyte count and decreases of IL-6, sCD163 and of sIL-2R (Figure 2). The IL-10/IL-6 ratio of serum (an index of the anti-inflammatory/pro-inflammatory balance in severe COVID-19^7^) was inversely associated with the absolute increase of the SOFA score on day 14 among anakinra-treated patients, compatible with the anti-inflammatory effect of anakinra. Remarkably, suPAR was increased among anakinra-treated patients on day 7 from baseline. Thus anakinra is associated with protection against progression to SRF even when the suPAR signal indicates an unfavourable outcome. The function of PBMCs of patients was modulated among anakinra-treated patients. More precisely, the production of IL-1β and IL-10 on day 7 was greater among patients who did not develop SRF pointing towards a restoration of the ability of the PBMCs to adapt to balanced production of anti-inflammatory and pro-inflammatory cytokines. This was further corroborated with the positive association between the IL-10/IL-1β ratio of production from PBMCs on day 7 with the ratio of serum IL-10/IL-6 of the same day (Figure S2).

**Figure 2.**
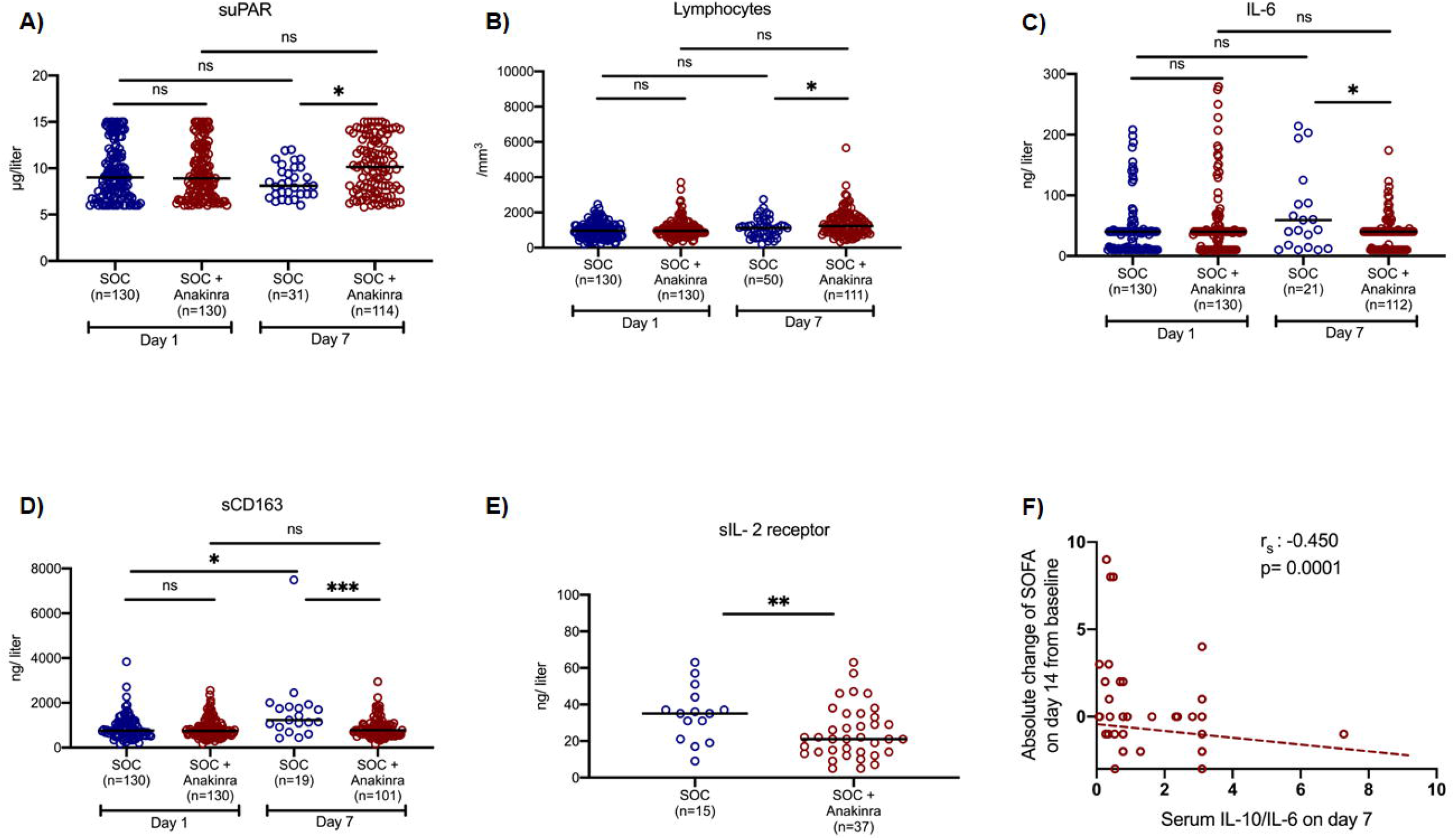
Effect of anakinra treatment on circulating mediators of patients with lower respiratory tract infection by SARS-CoV-2. Concentrations of A) suPAR (soluble urokinase plasminogen activator receptor); B) the absolute lymphocyte count; C) interleukin (IL)-6; and D) sCD163 of patients under standard-of-care treatment (SOC, blue colour) and of patients under SOC treatment with anakinra (red colour) at baseline (day of hospital admission) and day 7 of follow-up are shown. The concentrations of sIL-2R on day 7 are provided in panel E. The number of patients in each group is provided inside the bars. Lines refer to median values. Comparisons between different groups were performed by the Mann-Whitney U test and within the same group by the Wilcoxon test. Statistical comparisons: ns, non-significant; *P< 0.05; **P<0.01; ***P<0.0001 The correlation of the IL-10/IL-6 ratio with the absolute change of the SOFA (sequential organ failure assessment) score on day 14 from the baseline is also shown (panel F). The Spearman rank of order correlation co-efficient and the P-value of the correlation is provided.

The exploratory endpoints were the stay in the ICU and in hospital and the cost of hospitalization. Patients who were eventually intubated and admitted in the ICU showed a shorter median ICU stay when they had received previous treatment with anakinra (Figure S3). The median overall hospital stay was 13 days for those that were treated with anakinra plus SOC, and 15 days with only SOC treatment (Figure S4). These differences were associated with significant decrease of the overall cost of hospitalization from median €2,398.4 among SOC treated comparators to €1,291.4 among anakinra-treated patients (Figure S5).

### Safety

The adverse events (AEs) and serious adverse events (SAEs) that were captured during the study period of 14 days are listed in Table 3. The incidence of the same events was depicted among SOC treated comparators. As shown in Table 3, the incidence of these events was not greater in the anakinra group than comparators, with the only exception of leukopenia having a trend to be higher in the anakinra group. SAEs were fewer among anakinra-treated patients.

**Table 3.** Adverse Events and Serious Adverse Events. The incidence of the same events was depicted among standard-of-care treated comparators.

|  | Standard-of-care (N=130) | Standard-of-care +<br>Anakinra (N=130) | OR (95% CI) | P Value |
| --- | --- | --- | --- | --- |
| At least one SAE by day 14, n (%) | 63 (60.6) | 32 (29.4) | 0.27 (0.15-0.48) | **5.0 x 10<sup>-6</sup>** |
| Extended hospitalization, n (%) | 63 (60.6) | 32 (29.4) | 0.27 (0.15-0.48) | **5.0 x 10<sup>-6</sup>** |
| Death, n (%) | 16 (12.3) | 6 (4.6) | 0.35 (0.13-0.91) | **0.043** |
| Shock, n (%) | 56 (43.1) | 27 (20.8) | 0.34 (0.20-0.59) | **1.6 x 10<sup>-5</sup>** |
| Acute kidney injury, n (%) | 37 (28.5) | 15 (11.5) | 0.30 (0.15-0.58) | **7.0 x 10<sup>-5</sup>** |
| Any bacterial infection, n (%) | 30 (23.1) | 9 (6.9) | 0.22 (0.10-0.49) | **3.3 x 10<sup>-5</sup>** |
| Thromboembolic event, n (%) | 5 (4.8) | 2 (1.6) | 0.33 (0.06-1.73) | 0.252 |
| Pulmonary edema, n (%) | 0 (0) | 1 (0) | NA | 1.00 |
| At least one AE by day 14, n (%) | 89 (85.6) | 85 (78.7) | 0.62 (0.31-1.27) | 0.213 |
| Gastrointestinal disturbances, n (%) | 9 (6.9) | 15 (11.5) | 1.74 (0.74-4.17) | 0.284 |
| Electrolyte derangements, n (%) | 41 (31.5) | 35 (26.9) | 0.80 (0.47-1.37) | 0.496 |
| Elevated liver function tests, n (%) | 51 (39.2) | 40 (30.8) | 0.69 (0.41-1.15) | 0.193 |
| Anemia, n (%) | 26 (20.0) | 22 (16.9) | 0.82 (0.44-1.53) | 0.632 |
| Leukopenia, n (%) | 3 (2.3) | 11 (8.5) | 3.91 (1.07-14.37) | 0.051 |
| Thrombopenia, n (%) | 7 (5.4) | 9 (6.9) | 1.31 (0.47-3.62) | 0.797 |
| Headache, (%) | 2 (1.5) | 4 (3.1) | 2.03 (0.37-11.29) | 0.684 |
| Allergic reaction, n (%) | 7 (5.4) | 4 (3.1) | 0.56 (0.16-1.95) | 0.540 |
| Any heart arrhythmia, n (%) | 22 (16.9) | 9 (6.9) | 0.37 (0.16-0.83) | **0.020** |

Abbreviations: AE: non-serious adverse event; CI: confidence interval; n: number; NA: non-available; OR: odds ratio; SAE serious adverse event

## DISCUSSION

Anakinra treatment of COVID-19 patients admitted with LRTI and suPAR concentrations greater or equal than 6μg/l was associated with a relative decrease of the incidence of SRF by 72%. Anakinra-treated patients who were admitted to the ICU had a shorter stay in the ICU than those who did not receive anakinra. So apparently the benefit of previous anakinra treatment remained. This is also reflected by the overall decrease of 30-day mortality. An important point of the SAVE study is the strategy to select for early anakinra treatment using the predictive biomarker suPAR. In previous studies in COVID-19 and in sepsis, this marker turned out to be able to predict the likelihood for unfavourable outcome^4, 5, 8, 9^.

Other studies have reported favourable effects of anakinra treatment in COVID-19 pneumonia. In a retrospective analysis by Cavalli et al, 29 patients with respiratory failure and respiratory ratio below 100 mmHg were treated with high dose anakinra intravenously (5mg/kg twice daily). Treatment was associated with clinical improvement in 72% and remarkably higher survival rate^10^. In the Ana-COVID prospective study, 52 patients with confirmed COVID-19 and bilateral lung infiltrates and oxygen saturation less than 93% were treated with 100mg anakinra subcutaneously twice daily for three days followed by 100 mg subcutaneously once daily for another seven days. The study had a composite endpoint much similar to the endpoint of the SAVE study; i.e. MV and/or death. Anakinra treatment achieved a 78% decrease of this composite endpoint compared to 44 historical controls. However, studied comparators were not matched for co-administered medication such as azithromycin and hydroxychloqoquine^11^. In a retrospective study, 12 patients with COVID pneumonia and increased C-reactive protein were intravenously treated with anakinra 300 mg/day; none died^12^.

It is difficult to compare these three studies as enrolled patients had different stages of COVID and variable disease severity. Hence the clinical dilemma is which patients would benefit from anakinra, in what stage of COVID, and in what dosage regimen. In this respect, the SAVE study gives guidance: the best candidates are patients with high likelihood of SRF as defined by increased suPAR; anakinra presented one well-acceptable safety profile at the standard subcutaneous daily dose of 100 mg.

An important additional finding of the SAVE trial is that it provides mechanistic insight into the biological effects of anakinra: the treatment was associated with a reset of the pro-versus anti-inflammatory balance of the host. The production capacity of PMBCs for the anti-inflammatory IL-10 was increased and this was reflected by the serum IL-10/IL-6 ratio. IL-10/IL-6 is an index of the anti-inflammatory/pro-inflammatory balance which is severely disturbed in severe COVID-19, much more than in bacterial sepsis^7^. The described reset in the production capacity of the PBMCs was linked to clinical benefit since the serum IL-10/IL-6 ratio was inversely associated with the absolute change of SOFA score. Anakinra treatment also decreased the elevated serum concentrations of sCD163 and sIL2-R that are biomarkers of macrophage activation^13, 14^. It was been noted before that patients with COVID-19 who deteriorate have characteristics of macrophage activation^15^; the decrease of these biomarkers indicates attenuation of macrophage activation among anakinra-treated patients.

The SAVE trial was designed in mid-March 2020 at the beginning of the pandemic in Greece. It was chosen to adapt an open-label and single-arm design in an attempt to help as many patients as possible since no SOC was framed at that time period. The SOC parallel comparators were optimally matched without differences in baseline severity and co-administered treatment. The extremely low P value of significance between patients receiving anakinra with SOC treatment and patients receiving only SOC treatment is favoring the importance of the findings. When the study was started, dexamethasone treatment was not yet part of the SOC treatment but in the following months many patients received dexamethasone. Since dexamethasone acts as a cytokine-inhibiting agent, it is an important question how strong would the effect of anakinra be when patients also receive dexamethasone. Although this is not a preset endpoint, post-hoc analysis shows that anakinra is also protective in patients receiving dexamethasone (Table S2)

The impact of this interim analysis in the Greek Infectious Diseases community was immense. Enthusiasm shared among physicians led to high demand for joining the trial from many other study sites. As such, the SAVE on-going trial has been amended for the inclusion of 27 study sites.

In conclusion, we propose a novel strategy using suPAR as an early biomarker that can effectively identify those patients at high risk for SRF. In these patients, prophylactic treatment with regular doses of anakinra is associated with prevention of the incidence of SRF. The restoration of the pro-inflammatory/anti-inflammatory balance is proposed as the mechanism of anakinra action.

## Data Availability

All data are available from the senior author (E. J Giamarellos-Bourboulis) upon request

## Funding and disclosure

The study is supported in part by the Hellenic Institute for the Study of Sepsis, in part by Technomar Shipping Inc, in part by Swedish Orphan Biovitrum AB and in part by the Horizon 2020 grant RISKinCOVID.

Periklis Panagopoulos has received honoraria from GILEAD Sciences, Janssen, and MSD.

George N. Dalekos has acted as Advisor/Lecturer for Abbvie, Bristol-Myers Squibb, Gilead, Novartis, Roche, Amgen, MSD, Janssen, Ipsen and Pfizer, has received Grant support from Bristol-Myers Squib, Gilead, Roche, Janssen, Abbvie and Bayer and was or is currently PI in National & International Protocols sponsored by Abbvie, Bristol-Myers Squibb, Novartis, Gilead, Novo Nordisk, Genkyotex, Regulus Therapeutics Inc, Tiziana Life Sciences, Bayer, Astellas, Ipsen, Pfizer and Roche. G. Poulakou has received independent educational grants from Pfizer, MSD, Angelini, and Biorad.

Haralampos Milionis reports receiving honoraria, consulting fees and non-financial support from healthcare companies, including Amgen, Angelini, Bayer, Mylan, MSD, Pfizer, and Servier.

Jesper Eugen-Olsen is a cofounder, shareholder and CSO of ViroGates A7S, Denmark and is named inventor on patents on suPAR owned by Copenhagen University Hospital Hvidovre, Denmark.

Mihai G. Netea is supported by an ERC Advanced Grant (#833247) and a Spinoza grant of the Netherlands Organization for Scientific Research. He has also received independent educational grants from TTxD, GSK and ViiV HealthCare.

E.J. Giamarellos-Bourboulis has received honoraria from Abbott CH, Angelini Italy, InflaRx GmbH, MSD Greece, XBiotech Inc., and B·R·A·H·M·S GmbH (Thermo Fisher Scientific); independent educational grants from AbbVie Inc, Abbott CH, Astellas Pharma Europe, AxisShield, bioMérieux Inc, Novartis, InflaRx GmbH, and XBiotech Inc; and funding from the FrameWork 7 program HemoSpec (granted to the National and Kapodistrian University of Athens), the Horizon2020 Marie-Curie Project European Sepsis Academy (granted to the National and Kapodistrian University of Athens), and the Horizon 2020 European Grant ImmunoSep (granted to the Hellenic Institute for the Study of Sepsis).

The other authors do not report any conflict of interest.

The authors would like to thank the patients, families, clinical, laboratory and research staff who contributed to the trial.

