## SUPPLEMENTARY APPENDIX for "Anakinra To Prevent Respiratory Failure In COVID-19"

**Table of contents**

| **Content** | **Page** |
| --- | --- |
| Laboratory methods | 4 |
| The score of respiratory symptoms | 5 |
| Cost estimation | 6 |
| Reporting of Adverse events (AEs) and Serious Adverse Events (SAEs) | 6 |
| Supplementary Figures | 9 |
| Supplementary Figure 1. Trial profile and selection of standard-of-care comparators | 9 |
| Supplementary Figure 2. Effect of anakinra treatment on cytokine production capacity from peripheral blood mononuclear cells (PBMCs) | 10 |
| Supplementary Figure 3. Time until discharge from the intensive care unit (ICU) | 12 |
| Supplementary Figure 4. Time until hospital discharge. | 13 |
| Supplementary Figure 5. Cost of hospitalization. | 14 |
| Supplementary Tables | 15 |
| Supplementary Table 1. Anakinra as an Independent Protective Factor from Development of Severe Respiratory Failure (SRF) by Day 14 | 15 |
| Supplementary Table 2. Anakinra as an Independent Protective Factor from Development of Severe Respiratory Failure (SRF) by Day 14 among patients treated with dexamethasone | 17 |
| Supplementary Table 3. Anakinra as an Independent Protective Factor from 30-Day Mortality | 18 |

**LABORATORY METHODS**

Peripheral blood mononuclear cells (PBMCs) were isolated after gradient centrifugation over Ficoll (Biochrom, Berlin, Germany) for 20 minutes at 1400g. After three washings in ice-cold PBS pH 7.2, PBMCs were counted in a Neubauer plate with trypan blue exclusion of dead cells. They were then diluted in RPMI 1640 enriched with 2mM of L-glutamine, 500 μg/ml of gentamicin, 100 U/ml of penicillin G, 10 mM of pyruvate, 10% fetal bovine serum (Biochrom) and suspended in wells of a 96-well plate. The final volume per well was 200μl with a density of 2 x10^6^ cells/ml. PBMCs were exposed in duplicate for 24 hours or 5 days at 37^0^C in 5% CO_2_ to different stimuli: 10 ng/ml of *Escherichia coli* O55:B5 lipopolysaccharide (LPS, Sigma, St. Louis, USA) or 5x10^5^ colony forming units of heat-killed *Candida albicans*. Following incubation, cells were removed and analysed for flow cytometry. Concentrations of IL-1β, IL-6 and IL-10 were measured in cell supernatants or serum in duplicate by an enzyme immunoassay (Invitrogen, Carlsbad, California, USA). The lowest detections limits were: for IL-1β 10 pg/ml; for IL-6 10 pg/ml; and for IL-10 5 pg/ml. Concentrations of ferritin (ORGENTEC Diagnostika GmbH, Mainz, Germany), sCD163 (Affymetrix Inc, Santa Clara, CA) and sIL-2R (ORGENTEC Diagnostika GmbH, Mainz, Germany) were measured in serum by an enzyme-immunoassay; the lower limit of detection was 75ng/ml for ferritin; 0.31 ng/ml for sCD163; and 0.5 ng/ml for sIL-2R.

**THE SCORE OF RESPIRATORY SYMPTOMS**

| **Symptom** | **Absent (score=0)** | **Mild (score=1)** | **Moderate (score=2)** | **Severe (score=3)** |
| --- | --- | --- | --- | --- |
| **Cough** | No cough or resolution | Cough present but does not interfere with subject’s usual daily activities | Cough present, frequent and does interfere with some of the subject’s usual daily activities | Cough present throughout day and night; limits most of the subjects’ usual daily activities and sleep patterns |
| **Chest pain** | No chest pain or resolution of chest pain | Chest pain present occasionally with deep breathing but does not interfere with subject’s usual daily activities | Chest pain present with normal breaths and does interfere with the subject’s usual daily activities | Chest pain present at rest and/or with shallow breathing; limits most of the subject’s usual daily activities |
| **Shortness of breath/ dyspnea** | No shortness of breath or resolution | Shortness of breath with strenuous activities only but does not interfere with subject’s usual daily activities | Shortness of breath with usual activities and does interfere with the subject’s usual daily activities | Shortness of breath with minimal exertion or at rest; limits most of the subject’s usual daily activities |
| **Sputum** | No coughing up of phlegm/sputum or resolution | Subject coughs up a small amount of phlegm/sputum | Subject coughs up a moderate amount of phlegm/sputum | Subject coughs up a large amount of phlegm/sputum |

### **COST ESTIMATION**

### Hospitalization cost was calculated per patient in Euros as the sum of all administered medicines and the addition of the nominal cost of daily stay in the intensive care unit or in the general ward. The unit price for counted items derived from the official pricelist as defined by the Greek government (KYA 4^α^/οικ.13740/27.03.2012; government gajette 4898 τΒ/01.1.2018). The cost of human resources (salaries of nursing and medical personnel) was not counted. Comparisons were done by the Mann-Whitney U test.

### **REPORTING OF ADVERSE EVENTS**

Adverse events (AEs) and Serious Adverse Events (SAEs) were collected from baseline until the last patient’s evaluation per protocol. Investigators were monitoring subjects for adverse events and were responsible for recording ALL AEs/SAEs occurring to a patient during the trial.

An adverse event was defined as any undesirable medical occurrence in a subject administered a pharmaceutical product and which did not necessarily have a causal relationship with this treatment. The time relationship was defined from the moment the AE occurs during therapeutic treatment until 5 half-lives after treatment discontinuation. The adverse event may be a sign, a symptom, or an abnormal laboratory finding.

If an adverse event met any of the following criteria, it was considered SAE:

- **Life-threatening situation:** The subject was at risk of death at the time of the adverse event/experience. It does not refer to the hypothetical risk of death.
- **Inpatient hospitalization** or prolongation of existing hospitalization.
- **Persistent or significant disability/incapacity:** This was not intended to include transient interruption of daily activities.
- **Congenital anomaly/birth defects:** Any structural abnormality in subject’s offspring that occurred after intrauterine exposure to treatment.
- **Important medical events/experiences** that may not result in death, be life-threatening, or require hospitalization may be considered a serious adverse event/experience when, based upon appropriate medical judgment, **they may jeopardize the subject and may require medical or surgical intervention to prevent one of the outcomes listed above,** i.e., death, a life-threatening adverse event/experience, inpatient hospitalization or prolongation of existing hospitalization, a persistent or significant disability/incapacity, or a congenital anomaly/birth defect.
- **Spontaneous and elective abortions** experienced by study subject.

**A non-serious adverse event** was any untoward medical occurrence in a patient or subject who is administered a pharmaceutical product, and which does not necessarily have a causal relationship with this treatment. A non-serious adverse event is one that does not meet the definition of a serious adverse event given above.

*Relationship to the drug*

The following definitions were used to assess the relationship of the adverse event to study drug:

- **Probably Related**: The adverse event had a strong time relationship to the drug or relapses if re-induced; another etiology is improbable or clearly less probable.
- **Possibly Related**: The adverse event had a strong time relationship to the drug; alternative aetiology is as probable or less probable.
- **Probably not Related**: The adverse event had a slight or no time relationship to the drug; there is a more probable alternative aetiology.
- **Unrelated**: The adverse event was due to an underlying or concomitant disease or to another pharmaceutical product and is not related to the drug (no time relationship and a much more probable alternative aetiology).

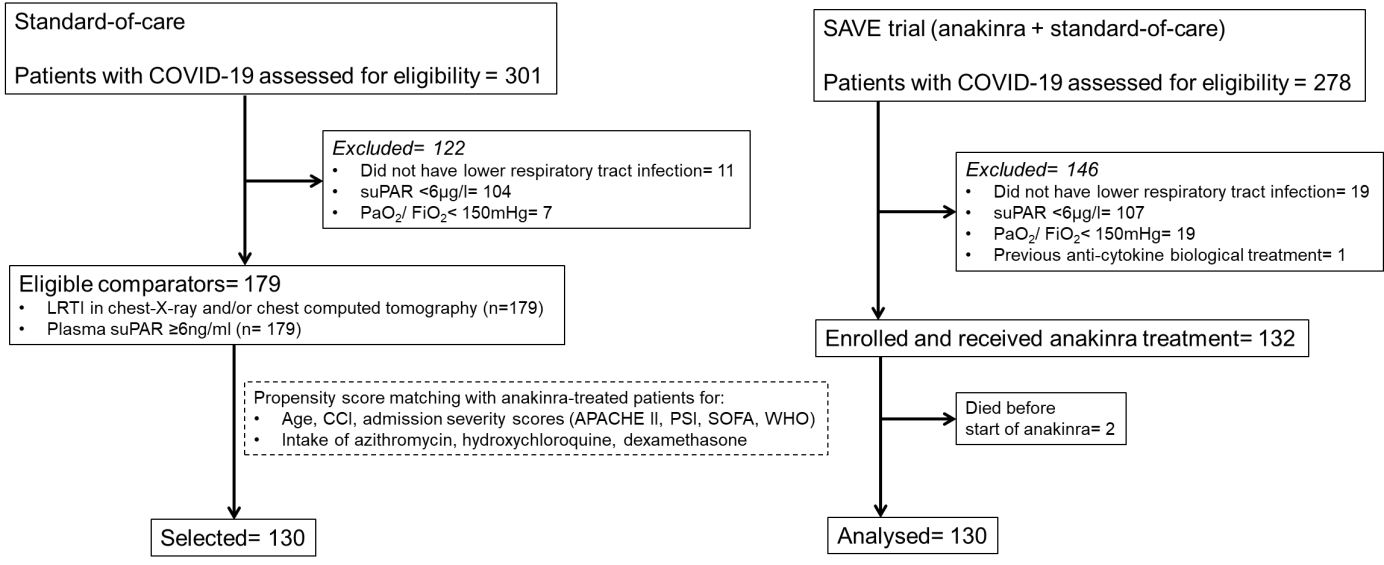

**Figure S1 Trial profile and selection of standard-of-care comparators**

Abbreviations APACHE II: acute physiology and chronic health evaluation; CCI: Charlson’s comorbidity index; COVID-19: infection by the new coronavirus SARS-CoV-2; LRTI: lower respiratory tract infection; PaO_2_/FiO_2_: respiratory fraction of partial oxygen pressure to fraction of inspired oxygen; PSI: pneumonia severity index; SOFA: sequential organ failure assessment; suPAR: soluble urokinase plasminogen activator receptor; WHO: severity classification of COVID-19 by the World Health organization

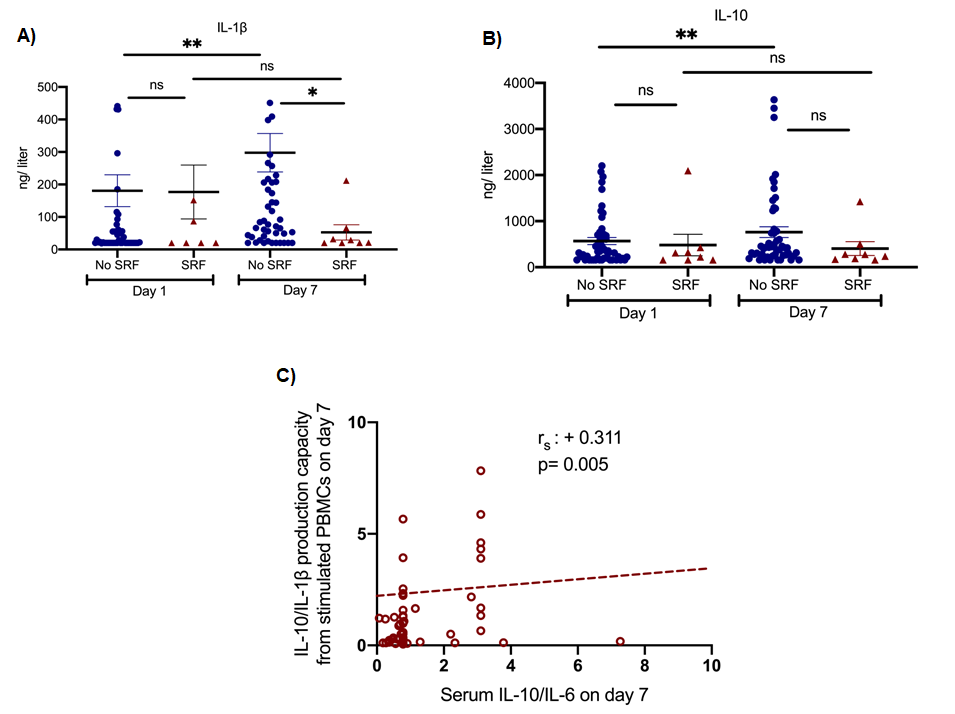

**Figure S2 Effect of anakinra treatment on cytokine production capacity from peripheral blood mononuclear cells (PBMCs)**

PBMCs were isolated before and seven days after start of treatment with anakinra. PBMCs were stimulated with lipopolysaccharide (LPS) of *Escherichia coli* O55:B5 for the production of interleukin (IL)-1β and with heat-killed *Candida albicans* for the production of IL-10. Production of IL-1β (panel A) and of IL-10 (panel B) is presented separately for patients who developed by day 14 severe respiratory failure (SRF) or not.

Lines refer to median values. Comparisons between different groups were performed by the Mann-Whitney U test and within the same group by the Wilcoxon test.

Statistical comparisons: ns, non-significant; *P< 0.05; **P<0.01

The correlation between the ratio of IL-10/IL-1β production by PBMCs and of the ratio of IL-10/IL-6 in serum on day 7 is provided in panel C. The Spearman rank of order correlation co-efficient and the P-value of the correlation is provided.

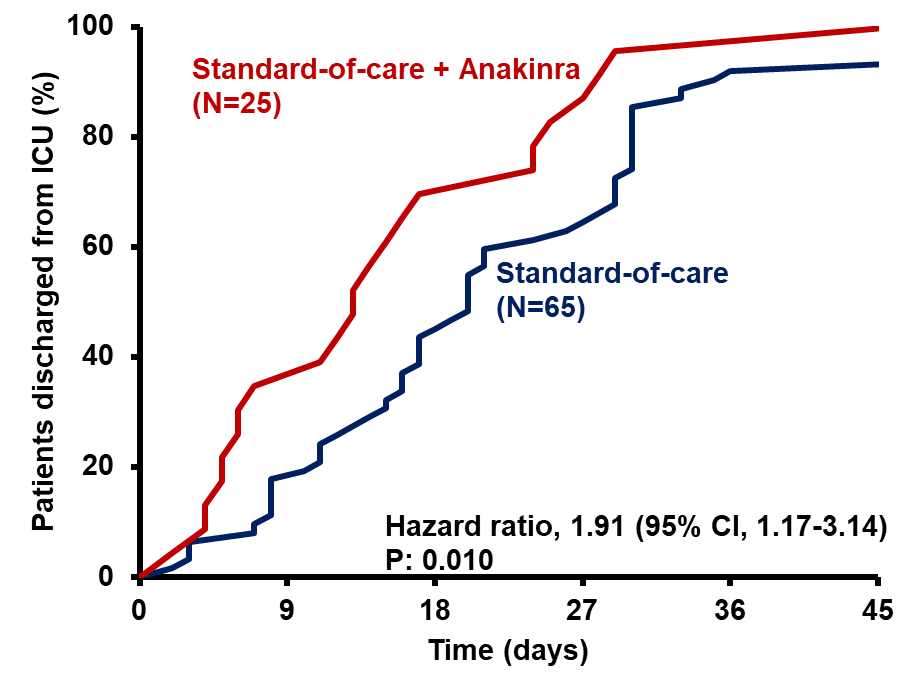

**Figure S3 Time until discharge from the intensive care unit (ICU)**

The analysis includes patients who were intubated and admitted in the ICU the first 14 days from start of anakinra (n= 25) or standard-of-care (n= 65).

CI: confidence interval

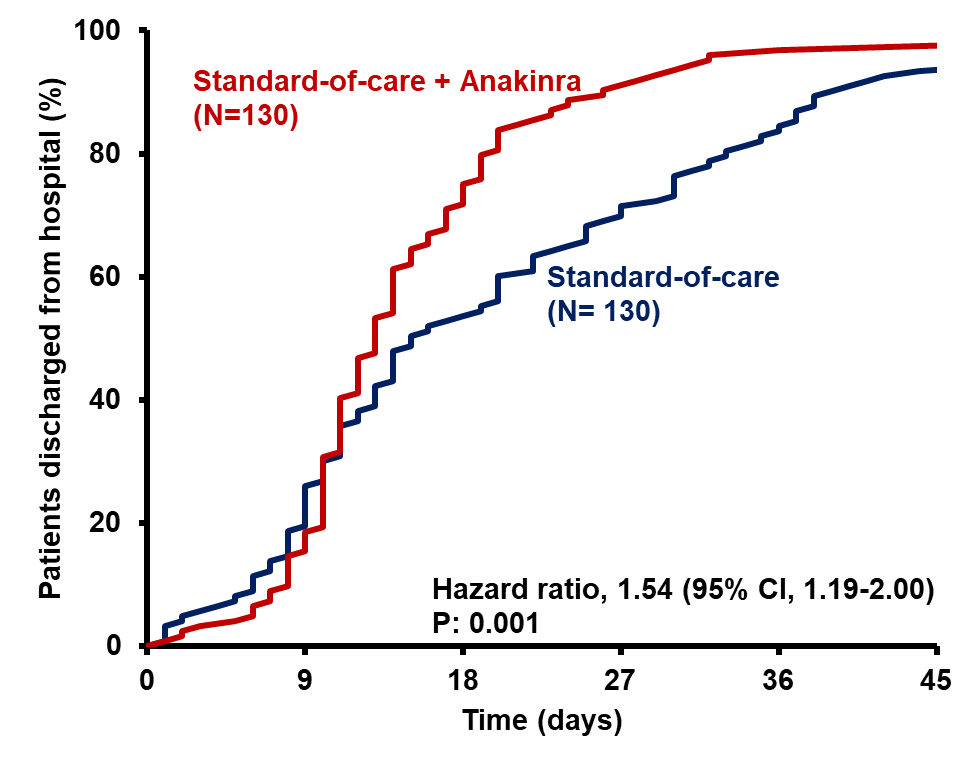

**Figure S4 Time until hospital discharge for all studied patients**

CI: confidence interval

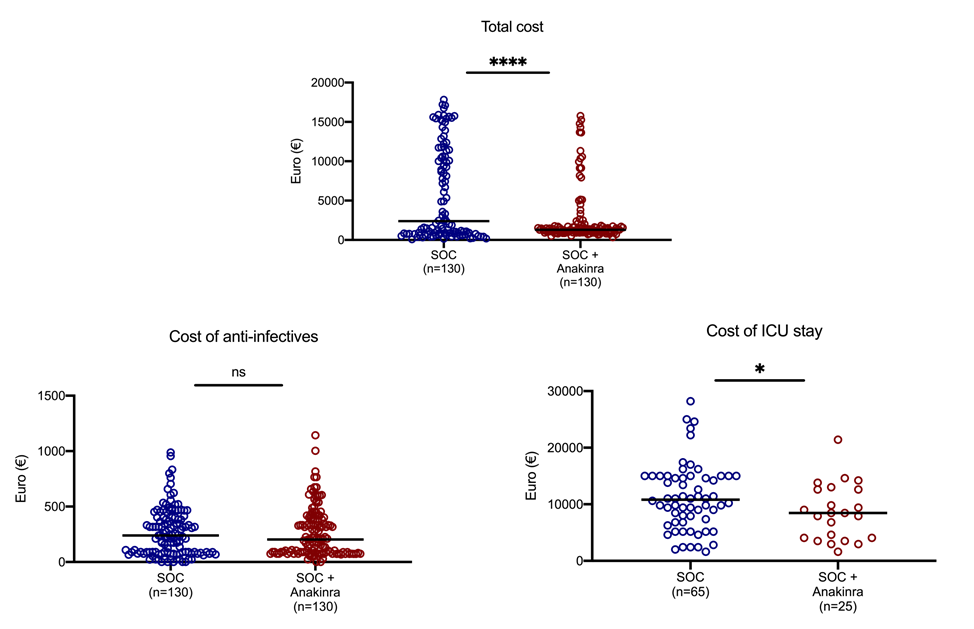

**Figure S5 Cost of hospitalization**

The three main categories of cost are shown: total cost; cost of anti-infectives; and cost of stay in the intensive care unit.

Statistical comparisons between groups ns: non-significant; *P<0.05; ****P<0.0001

**Table S1 Anakinra as an Independent Protective Factor from Development of Severe Respiratory Failure (SRF) by Day 14** Univariate and multivariate (Cox Forward Conditional) models, for patients receiving anakinra with Standard-of-care treatment (SOC) and for comparators receiving only SOC treatment are presented. Only admission variables that differ significantly between patients who developed and those who did not develop SRF by day 14 are provided. Results are provided after four steps of analysis.

|  |  |  | **Univariate analysis** | | **Multivariate analysis** | |
| --- | --- | --- | --- | --- | --- | --- |
| **Variable, no. (%)** | **SRF (-) (N=154)** | **SRF (+) (N=106)** | **HR (95% CI)** | **P- Value** | **HR (95% CI)** | **P- Value** |
| Anakinra treatment, n (%) | 101 (65.6) | 29 (27.4) | 0.30 (0.20-0.47) | **4.0 x 10^-8^** | 0.28 (0.18-0.44) | **2.4 x 10^-8^** |
| APACHE II, mean (SD) | 6.02 (3.24) | 7.87 (3.38) | 1.12 (1.07-1.19) | **2.0 x 10^-5^** |  |  |
| SOFA score, mean (SD) | 1.77 (1.11) | 2.70 (1.35) | 1.58 (1.38-1.82) | **8.1 x 10^-11^** | 1.41 (1.21-1.65) | **9.0 x 10^-5^** |
| Pneumonia Severity Index, mean (SD) | 66.5 (21.8) | 73.1 (19.2) | 1.01 (1.00-1.02) | **0.017** |  |  |
| Severe COVID-19 by WHO classification, n (%) | 65 (42.2) | 77 (72.6) | 2.78 (1.81=4.27) | **3.0 x 10^-5^** | 1.74 (1.09-2.79) | **0.020** |
| Soluble uPAR, μg per liter, median (Q_1_-Q_3_) | 8.3 (6.7-11.0) | 10.0 (8.0-13.9) | 1.10 (1.04-1.17) | **0.001** | 1.07 (1.01-1.14) | **0.022** |
| Lymphocytes /mm^3^, median (Q_1_-Q_3_) | 1,000 (733-1,315) | 930 (579-1,197) | 0.99 (0.99-1.00) | **0.026** |  |  |
| C-reactive protein, mg per liter, median (Q_1_-Q_3_) | 41.1 (12.7-95.2) | 73.4 (30.6-153.1) | 1.00 (1.00-1.00) | **3.2 x 10^-4^** |  |  |
| Ferritin, μg per liter, median (Q_1_-Q_3_) | 525.0 (280.0-804.5) | 641.5 (451.3-1,556.8) | 1.00 (1.00-1.00) | **3.6 x 10^-7^** |  |  |
| PaO_2_/FiO_2_, mmHg, median (Q_1_-Q_3_) | 330.8 (231.9-386.7) | 244.4 (161.7-305.8) | 0.99 (0.99-0.99) | **2.6 x 10^-8^** |  |  |
| Treatment with 3^rd^ generation cephalosporin, n (%) | 73 (47.4) | 30 (28.3) | 0.53 (0.35-0.82) | **0.004** |  |  |
| Treatment with piperacillin/tazobactam, n (%) | 37 (24.0) | 42 (39.6) | 1.60 (1.08-2.38) | **0.019** |  |  |
| Treatment with carbapenem, n (%) | 7 (4.5) | 19 (17.9) | 2.79 (1.69-4.60) | **5.9 x 10^-5^** | 2.19 (1.26-3.83) | **0.006** |
| Treatment with glycopeptide, n (%) | 1 (0.6) | 6 (5.7) | 2.60 (1.14-5.95) | **0.023** |  |  |
| Treatment with dexamethasone, n (%) | 46 (29.9) | 53 (50.0) | 1.73 (1.19-2.54) | **0.005** |  |  |

Abbreviations: APACHE: acute physiology and chronic health evaluation; CI: confidence interval; HR: hazard ratio; n: number; PaO_2_/FiO_2_: ratio of partial oxygen pressure to the fraction of inspired oxygen; Q: quartile; SD: standard deviation; SOFA: sequential organ failure assessment; SRF: severe respiratory failure; uPAR: urokinase-type plasminogen activator receptor.

**Table S2 Anakinra as an Independent Protective Factor from Development of Severe Respiratory Failure (SRF) by Day 14 among patients treated with dexamethasone** Univariate and multivariate (Cox Forward Conditional) models, for the Anakinra with Standard-of-care group (SOC) and SOC comparators are presented. Only admission variables that differ significantly between patients who developed and those who did not develop SRF by day 14 are provided.

|  |  |  | **Univariate analysis** | | **Multivariate analysis** | |
| --- | --- | --- | --- | --- | --- | --- |
| **Variable no. (%)** | **SRF (-) (N=46)** | **SRF (+) (N=53)** | **HR (95% CI)** | **P- Value** | **HR (95% CI)** | **P- Value** |
| Anakinra treatment, n (%) | 31 (67.4) | 21 (39.6) | 0.55 (0.32-0.97) | **0.038** | 0.56 (0.32-0.97) | **0.038** |
| Soluble uPAR, μg per liter, median (Q_1_-Q_3_) | 9.60 (7.2-12.6) | 11.6 (7.3-12.6) | 1.10 (1.00-1.20) | **0.030** | 1.97 (1.14-3.41) | **0.018** |
| Ferritin, μg per liter, median (Q_1_-Q_3_) | 593.0  (411.3-968.5) | 633.0  (412.0-968.0) | 1.00 (1.00-1.00) | **0.025** |  |  |

Abbreviations: CI: confidence interval; HR: hazard ratio; n: number; Q: quartile; SRF: severe respiratory failure; uPAR: urokinase-type plasminogen activator receptor.

**Table S3 Anakinra as an Independent Protective Factor from 30-Day Mortality.** Univariate and multivariate (Cox Forward Conditional) models, for the Anakinra with Standard-of-care (SOC) group and SOC comparators are presented. Only admission variables that differ significantly between patients who died and those who did not die are provided. Results are provided after four steps of analysis.

|  |  |  | **Univariate analysis** | | **Multivariate analysis** | |
| --- | --- | --- | --- | --- | --- | --- |
| **Parameters, no. (%)** | **Survival (N=216)** | **Death (N=44)** | **HR (95% CI)** | **P- Value** | **HR (95% CI)** | **P- Value** |
| Anakinra treatment, n(%) | 115 (53.2) | 15 (34.1) | 0.49 (0.26-0.91) | **0.024** | 0.49 (0.25-0.97) | **0.041** |
| Charlson’s Comorbidity Index, mean (SD) | 2.62 (1.98) | 3.86 (1.89) | 1.29 (1.13-1.48) | **2.0 x 10^-4^** | 1.26 (1.09-1.48) | **0.002** |
| APACHE II score, mean (SD) | 6.29 (3.31) | 9.14 (2.93) | 1.24 (1.14-1.35) | **2.7 x 10^-5^** |  |  |
| SOFA score, mean (SD) | 1.90 (1.15) | 3.34 (1.36) | 2.13 (1.73-2.63) | **2.1 x 10^-12^** | 2.03 (1.61-2.55) | **1.8 x 10^-9^** |
| Pneumonia Severity Index, mean (SD) | 66.7 (20.9) | 81.1 (17.6) | 3.66 (1.85-7.24) | **2.0 x 10^-4^** |  |  |
| Severe COVID-19 by WHO classification,  n (%) | 105 (48.6) | 37 (84.1) | 4.86 (2.17-10.92) | **1.2 x 10^-3^** |  |  |
| Soluble uPAR, μg per liter,  median (Q1-Q3) | 8.8 (6.8-11.4) | 10.4 (8.0-14.7) | 1.15 (1.05-1.26) | **0.003** |  |  |
| Lymphocytes/mm^3^, median (Q1-Q3) | 984 (699-1,275) | 839 (545-1,167) | 0.999 (0.99-1.00) | **0.024** |  |  |
| C-reactive protein, mg per liter,  median (Q1-Q3) | 50.7 (17.5-118.4) | 80.1 (18.7-156.7) | 1.003 (1.00-1.01) | **0.033** |  |  |
| PaO_2_/FiO_2_ mmHg, median (Q1-Q3) | 301 (224.6-374.4) | 195.8 (88.2-290.4) | 0.99 (0.98-0.99) | **3.4 x 10^-7^** |  |  |
| History of type 2 diabetes mellitus, n (%) | 53 (24.5) | 20 (45.5) | 2.38 (1.31-4.31) | **0.004** |  |  |
| Treatment with 3^rd^ generation cephalosporin, n (%) | 93 (43.1) | 10 (22.7) | 0.43 (0.21-0.88) | **0.020** |  |  |
| Treatment with piperacillin/tazobactam,  n (%) | 59 (27.3) | 20 (45.5) | 2.03 (1.11-3.69) | **0.021** |  |  |
| Treatment with remdesivir, n (%) | 11 (5.1) | 8 (18.6) | 3.22 (1.50-6.95) | **0.003** | 1.27 (1.09-1.47) | **0.002** |

Abbreviations: APACHE: acute physiology and chronic health evaluation; CI: confidence interval; HR: hazard ratio; n: number; PaO_2_/FiO_2_: ratio of partial oxygen pressure to the fraction of inspired oxygen; q: quartile; SD: standard deviation; SOFA: sequential organ failure assessment; uPAR: urokinase-type plasminogen activator receptor.
